# Nutrient Composition of Foods Represented in the U.S. Food and Nutrient Database for Dietary Studies, 2013-2023

**DOI:** 10.64898/2026.06.17.26355853

**Authors:** Omar Ihab Moussa, Moaz Elsayed Abouelmagd, Belal Mohamed Hamed, Asmaa Zakria Alnajjar, Abdelrahman Shata

## Abstract

**Background:** The U.S. Food and Nutrient Database for Dietary Studies (FNDDS) is updated across NHANES dietary cycles and is central to U.S. nutrition surveillance. However, multi-cycle food-code-level changes in nutrient composition have not been comprehensively characterized across the full WWEIA nutrient panel.

**Objective:** To characterize ten-year temporal patterns in nutrient composition across five FNDDS cycles, evaluate pandemic-period food-code compositional stability, and distinguish exploratory mean-level signals from distributional heterogeneity that may reflect reformulation, database coverage, or food-code definition changes.

**Methods:** We analyzed five consecutive FNDDS biennial releases: 2013-14, 2015-16, 2017-18, 2019-20, and 2021-23. Nutrient values were extracted from the public FNDDS/FoodData Central release files and standardized to per-100-g food-code-level records. Cycle midpoints, 2013.5, 2015.5, 2017.5, 2019.5, and 2022.0, served as the independent variable in an exploratory ordinary least squares (OLS) regression. Mann-Kendall testing assessed monotonic rank trends, Welch’s ANOVA assessed food-code-level distributional heterogeneity, and pairwise Welch comparisons with Cohen’s d summarized pre-pandemic, pandemic-period, and post-pandemic differences. Equivalence testing using TOST with +/-10% bounds was restricted to the 2019-20 versus 2021-23 stability comparison. OLS sensitivity analyses were repeated after excluding the structurally atypical 2017-18 cycle.

**Results:** Sixty-three nutrients were analyzed. Eight nutrients showed nominal OLS trends, p < 0.05, but none remained significant after Bonferroni correction. Mann-Kendall testing identified two nominal monotonic signals, and none after adjustment. Welch’s ANOVA detected cycle-level distributional differences for 61 of 63 nutrients at nominal p < 0.05 and 57 of 63 after adjustment. Pairwise pandemic-period analyses showed many adjusted differences when the pre-pandemic baseline was compared with 2019-20 or 2021-23, but standardized effects were small, with all absolute Cohen’s d values < 0.20. No nutrient differed after adjustment between 2019-20 and 2021-23, and 39 of 48 primary analytes met +/-10% TOST equivalence criteria for that comparison. Slope estimates were directionally stable after excluding 2017-18, but nominal significance status remained sensitive to the short time series.

**Conclusions:** FNDDS food composition varied across cycles, but there was no clear decade-long linear trend for most nutrients. The main signal was a possible increase in total PUFA and linoleic acid, which may reflect changes in fat quality. The 2021-23 cycle was very similar to 2019-20, suggesting no major post-pandemic shift in the foods represented. These findings should be interpreted as food-database signals, not as direct estimates of what people consumed.

## 1. Introduction

The nutritional quality of a population’s diet is influenced by both what people consume and the nutrient composition assigned to those foods in national databases. Over the past decade, the U.S. food environment has been shaped by regulatory, market, and macroeconomic forces that may alter product formulation and the nutrient values recorded in surveillance systems. Between 2013 and 2023, the U.S. Food and Drug Administration (FDA) finalized the removal of partially hydrogenated oils, the principal source of industrially produced trans fatty acids, mandated the first-ever disclosure of added sugars on the Nutrition Facts label, and updated the label’s reference values for several micronutrients [1,2]. Two successive editions of the Dietary Guidelines for Americans (2015-2020 and 2020-2025) reinforced guidance to replace saturated fats with unsaturated fats, reduce sodium intake, and increase intake of dietary fiber and underconsumed micronutrients such as vitamin D, calcium, and potassium [3]. These policy signals created incentives for food reformulation, although the extent to which such changes appear in national composition databases remains uncertain.

Superimposed on these policy-driven changes, the COVID-19 pandemic introduced an acute macroeconomic disruption beginning in early 2020. Supply-chain fragmentation, shifts from food service to retail purchasing, and altered consumer stockpiling behavior changed observed food purchasing and eating behaviors [4,5]. Prior work has documented pandemic-associated changes in dietary behaviors; however, whether FNDDS food-code-level nutrient composition showed a large contemporaneous discontinuity remains unclear [6,7]. Because FNDDS captures foods as reported and coded for NHANES, rather than population intake weights, database-level analyses must distinguish changes in represented food codes from changes in consumed dietary exposure.

Existing longitudinal analyses of U.S. food composition have often focused on selected nutrients of concern, such as sodium, saturated fat, or added sugars[8,9]. This leaves surveillance gaps across individual fatty acids, fat-soluble vitamins, folate chemical forms, carotenoids, and trace minerals, all of which may be affected by reformulation, fortification, analytical updates, or changes in food-code coverage [10,11]. The USDA Food and Nutrient Database for Dietary Studies (FNDDS) provides the nutrient composition values underpinning NHANES-derived national dietary intake estimates and is updated across dietary survey cycles [12]. FNDDS characterizes foods and mixed dishes as consumed and coded in dietary recalls, but unweighted analyses of FNDDS foods should not be interpreted as direct estimates of population intake. This study aimed to describe how the nutrient profiles of foods consumed in the United States changed from 2013 to 2023, identify nutrients that showed meaningful shifts, and assess whether the food supply remained broadly stable after the COVID-19 pandemic.

## 2. Methods

### 2.1 Data Sources

Nutrient composition data were obtained from five consecutive FNDDS releases publicly available through the USDA FoodData Central repository (https://fdc.nal.usda.gov): FNDDS 2013-14, 2015-16, 2017-18, 2019-20, and 2021-23 [12]. Each release provides per-100-g edible-portion nutrient values for foods and beverages as coded for the corresponding NHANES dietary recall cycle. The unit of analysis in this study was the FNDDS food code; analyses were unweighted by population consumption volume. The complete WWEIA panel analyzed here included 63 analytes organized across proximate nutrients, sugars, fatty acids, minerals, fat-soluble vitamins, water-soluble vitamins, folate forms, carotenoids, and bioactives. The 2018 USDA Standard Reference Legacy database served only as a plausibility reference for nutrient magnitude and was not used as an inferential comparator [13].

Nutrient Classification and Tiering Strategy. The 63-analyte panel was divided into 48 primary analytes and 15 supplementary analytes. Primary analytes comprised the major macronutrients, fat subtypes, common minerals, major vitamins, and frequently reported fatty-acid classes used for the main text summaries. Supplementary analytes comprised five carotenoids, three additional folate chemical forms, five sparse long-chain or low-concentration PUFA species, caffeine, and theobromine. This tiering was based on data completeness and distributional behavior, not biological importance. Main inferential summaries are reported in Table 1 and Figures 1-4, while complete analyte-level results are provided in Supplementary Tables S1-S9 and Supplementary Figures S1-S5. Added sugars (WWEIA nutrient 539) lacked reportable values in the 2013-14 and 2015-16 cycles and therefore were not treated as a full-window decade-scale trend.

**Figure 1.**
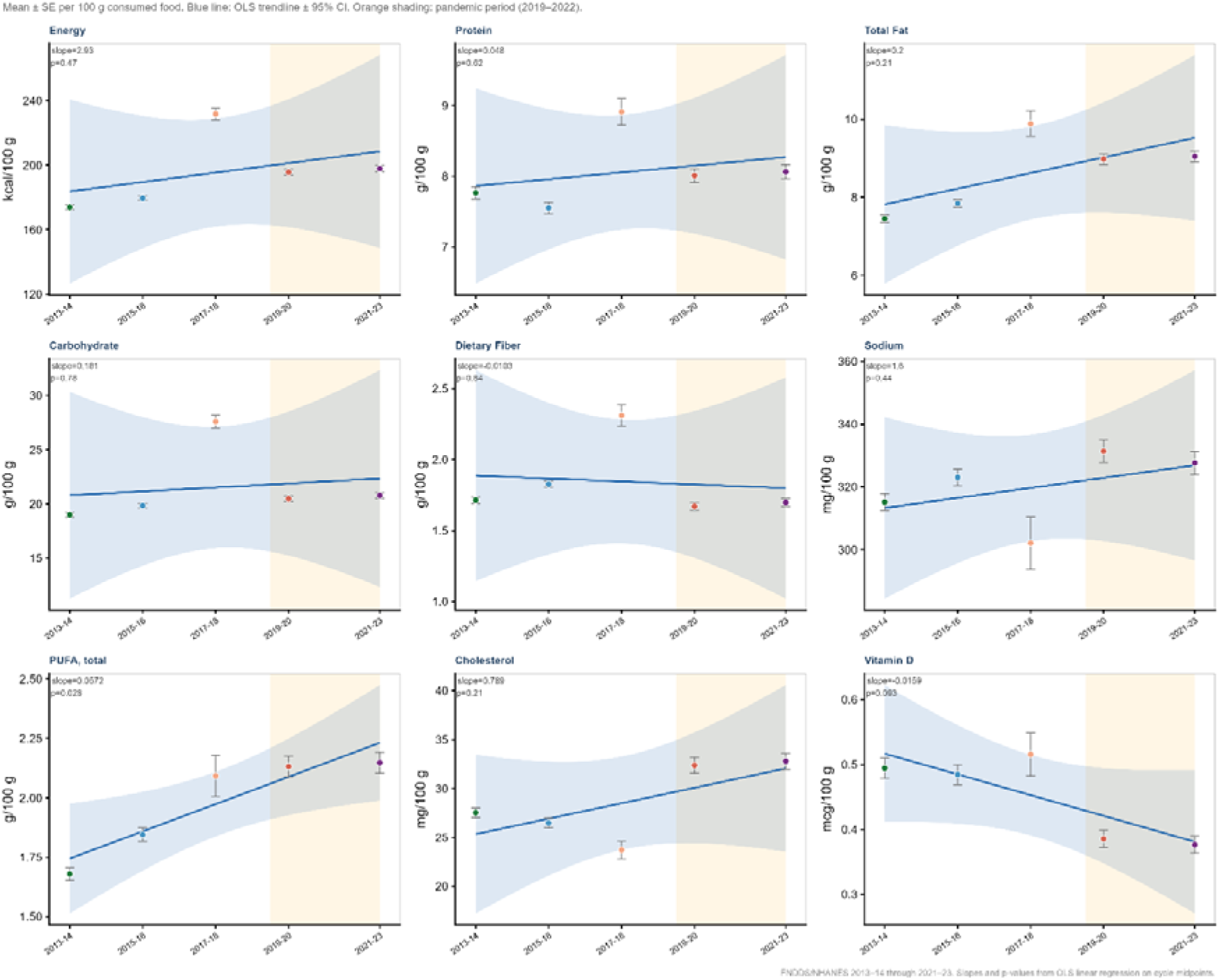
Temporal trends in selected nutrients across FNDDS cycles, 2013–2023.

**Table 1.**
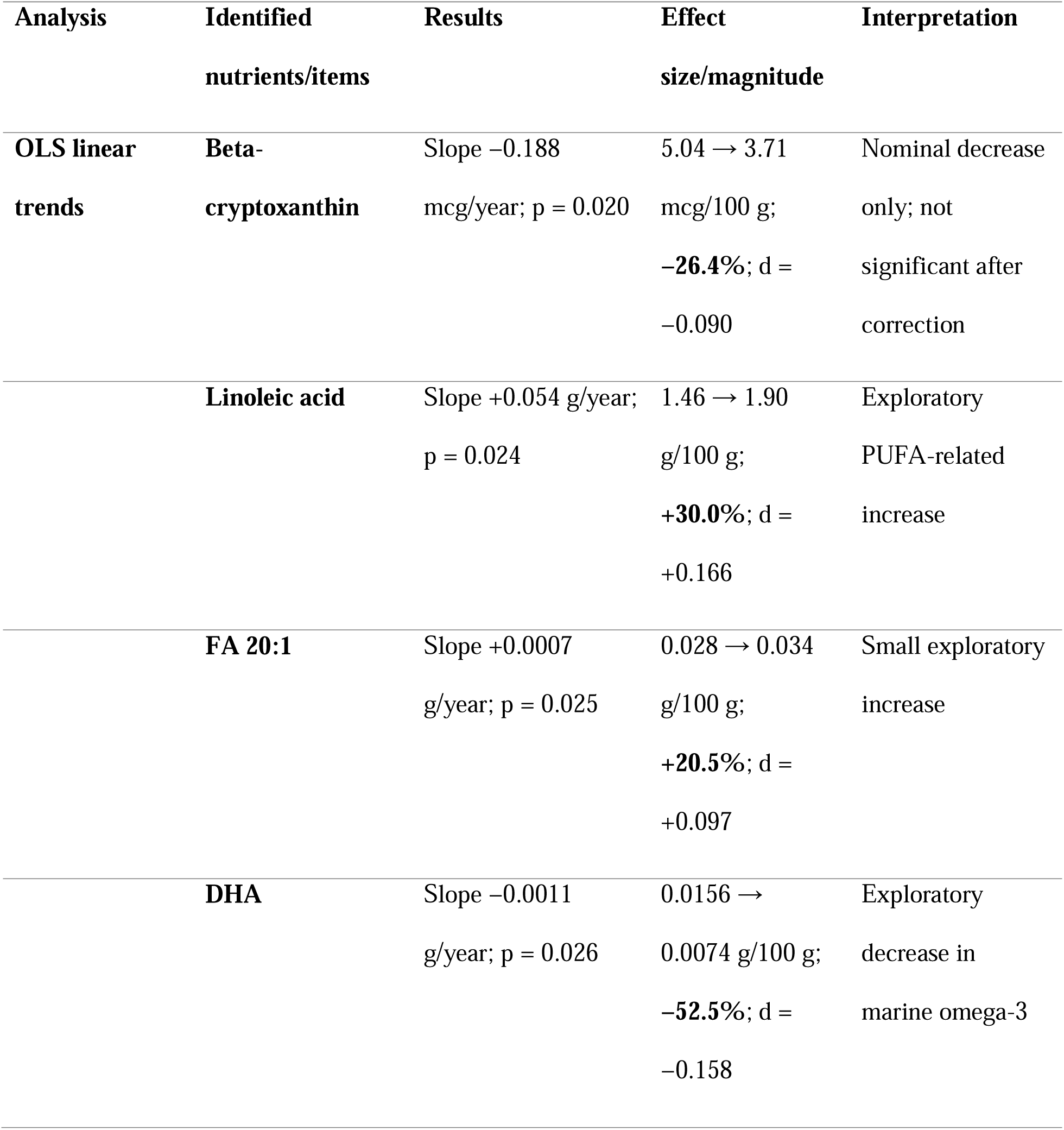
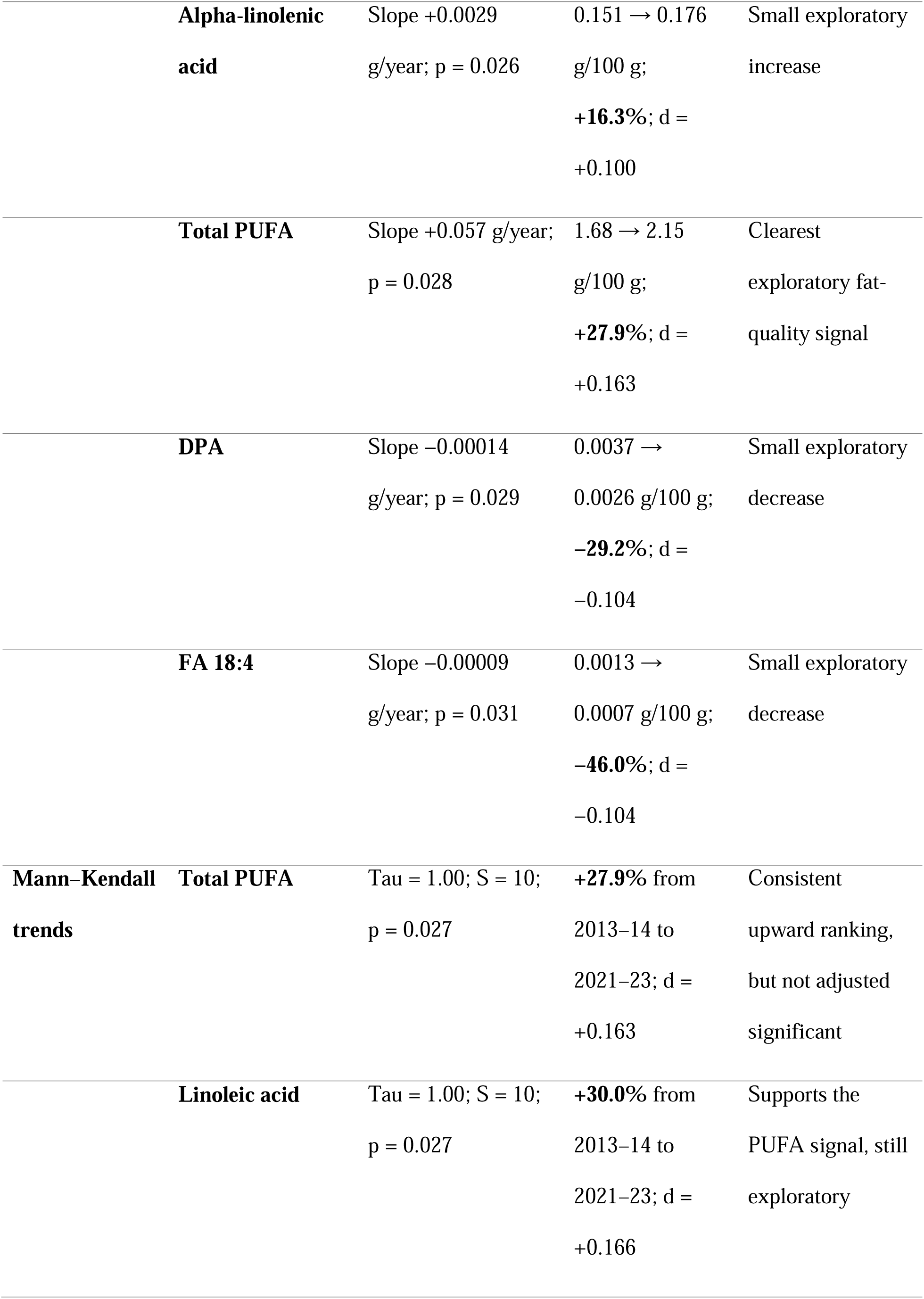
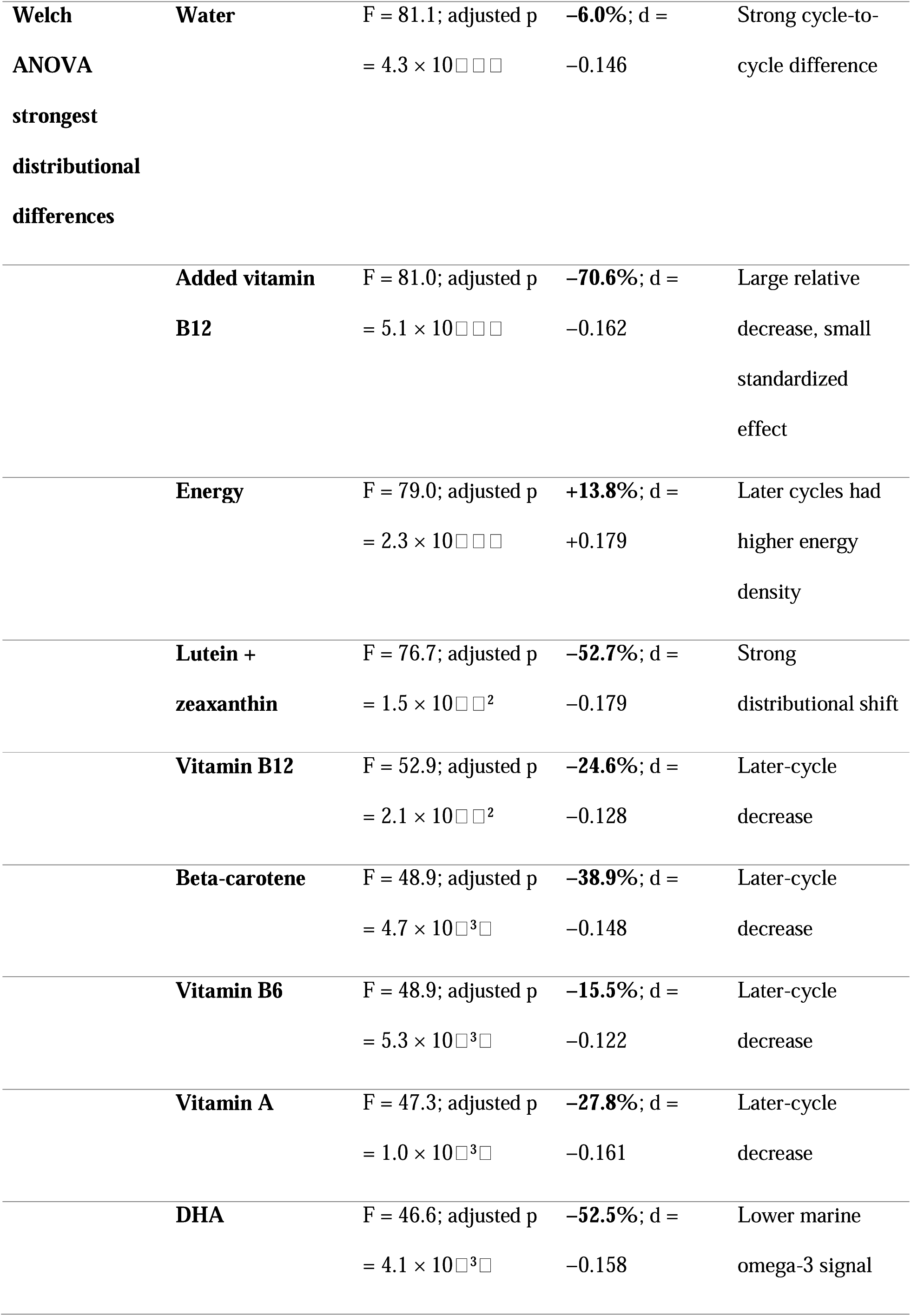
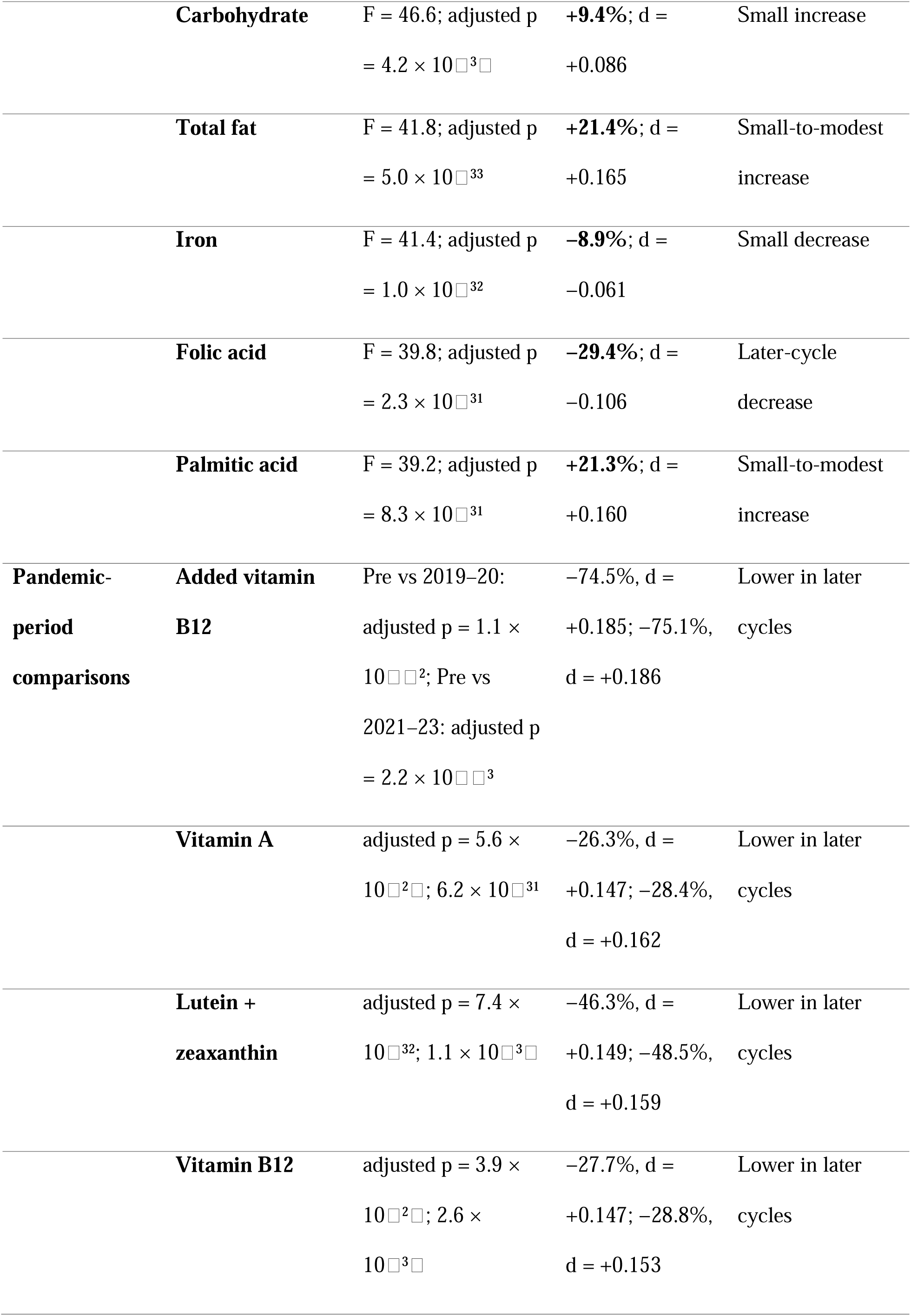
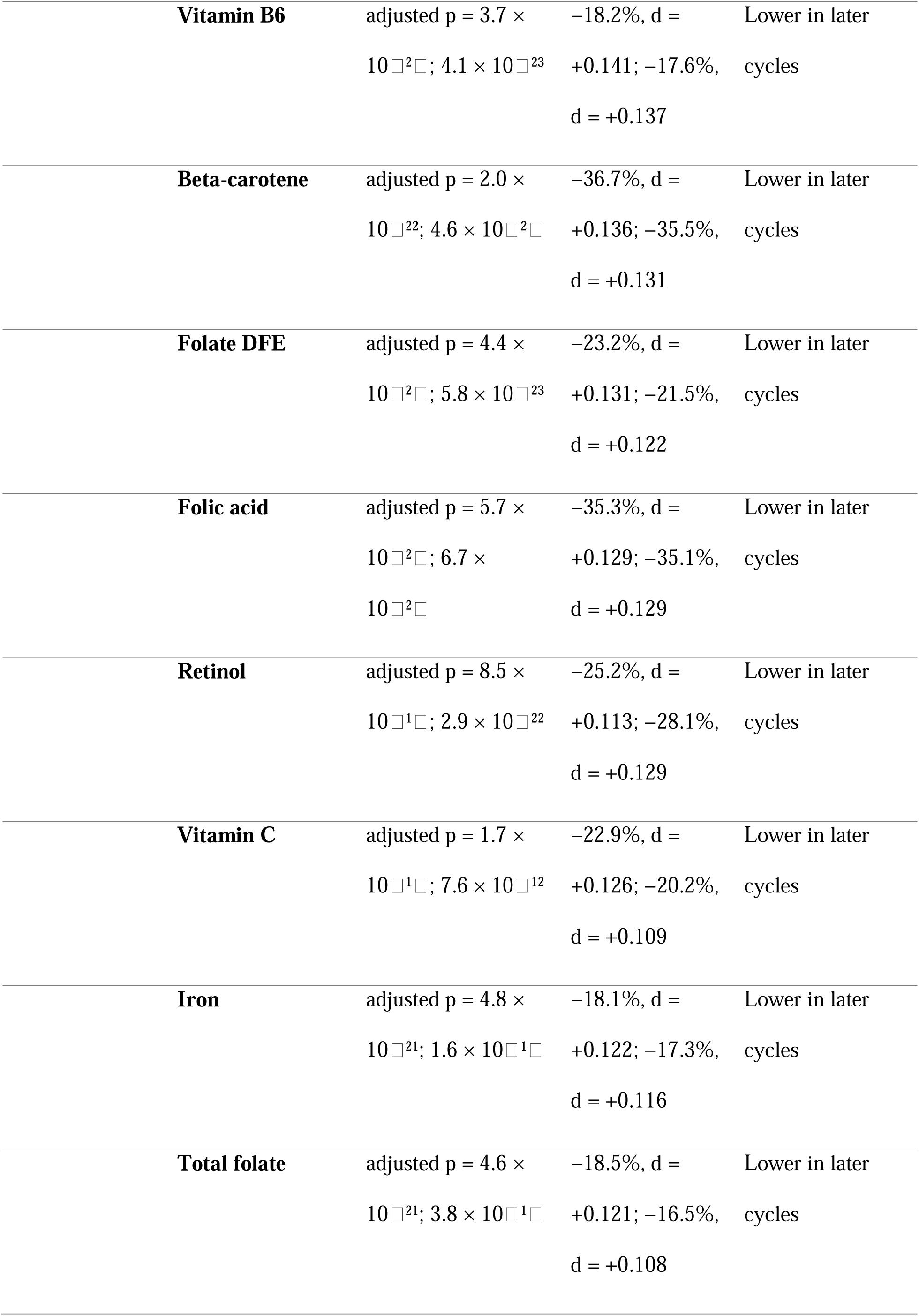
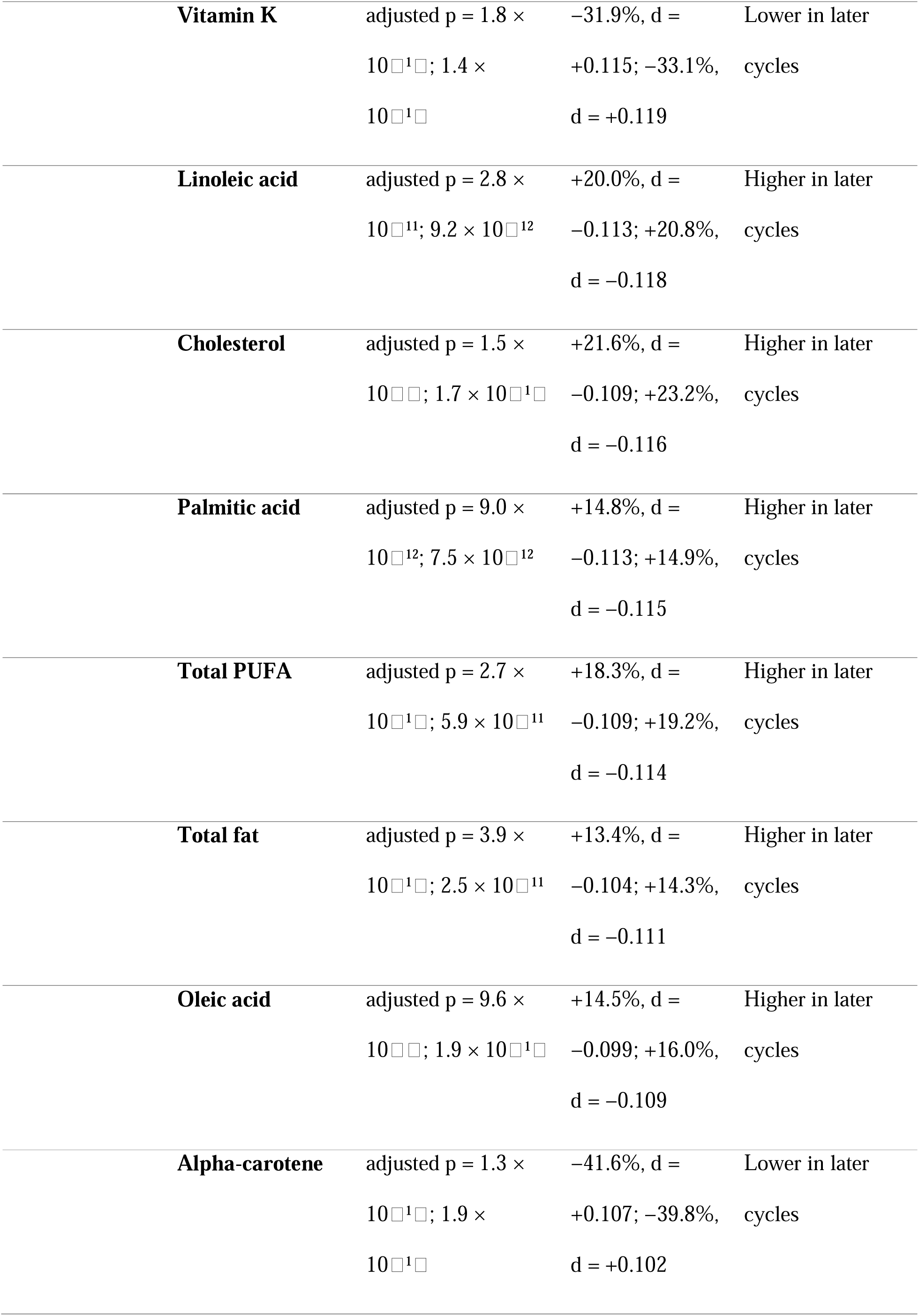
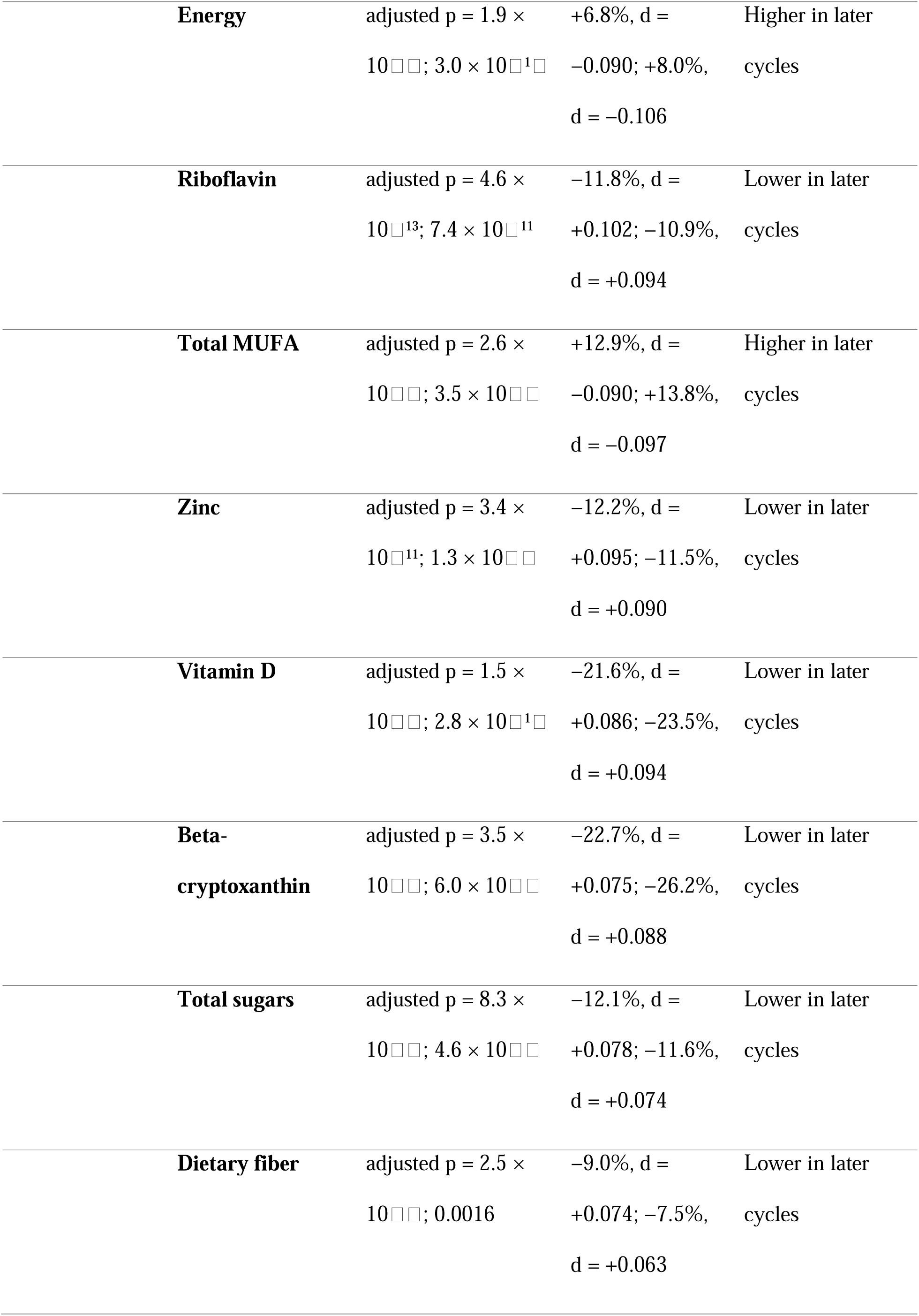
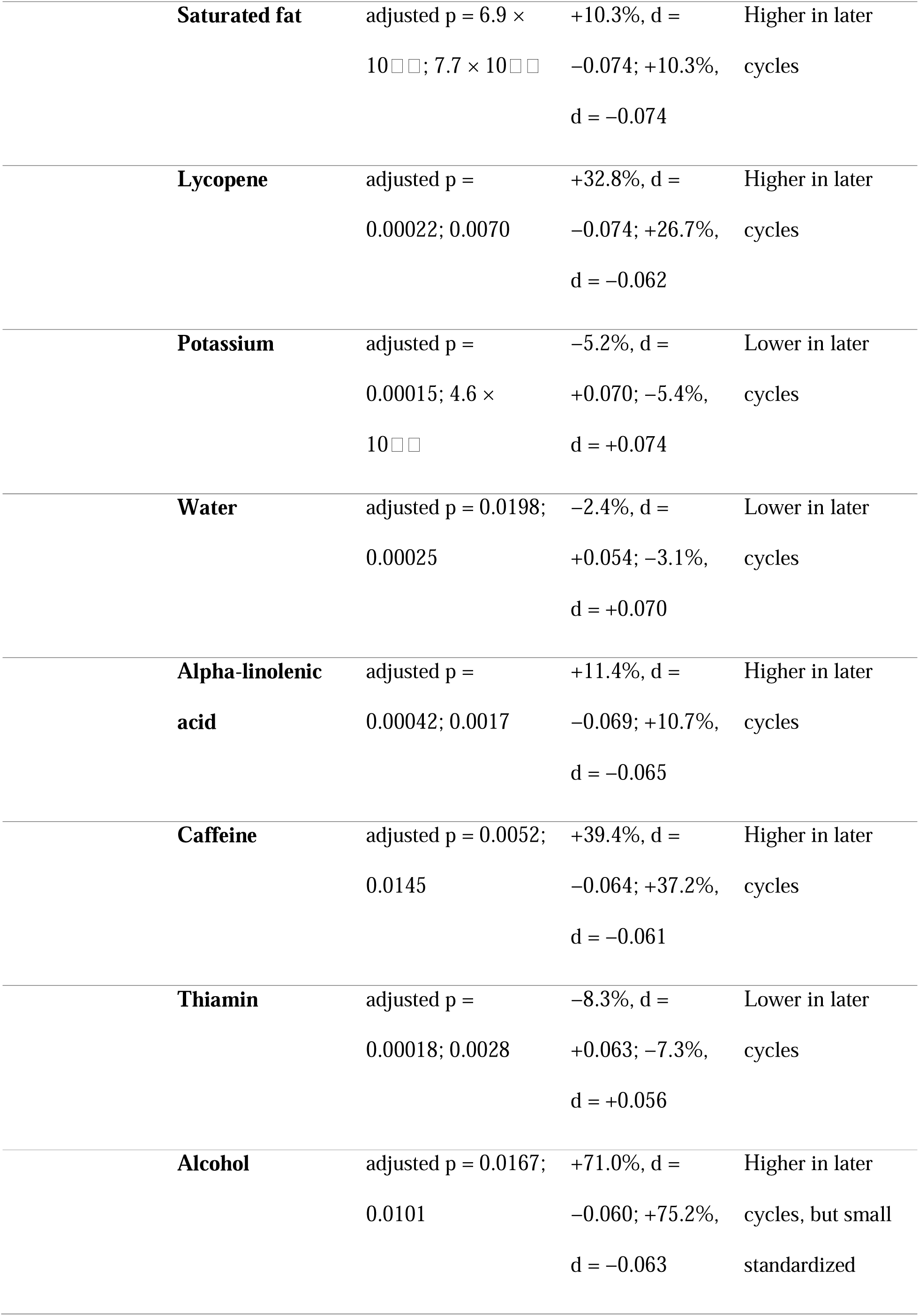
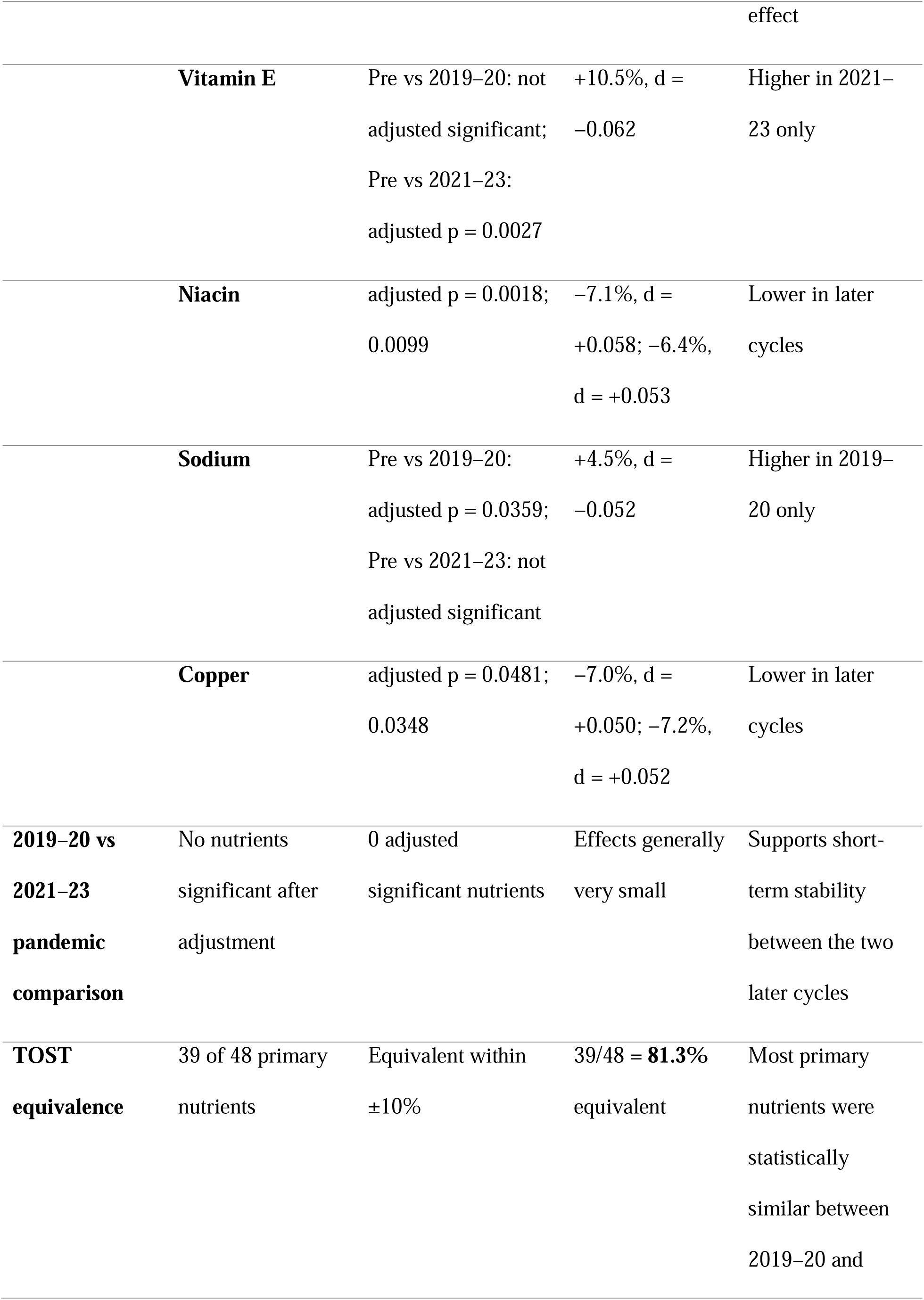
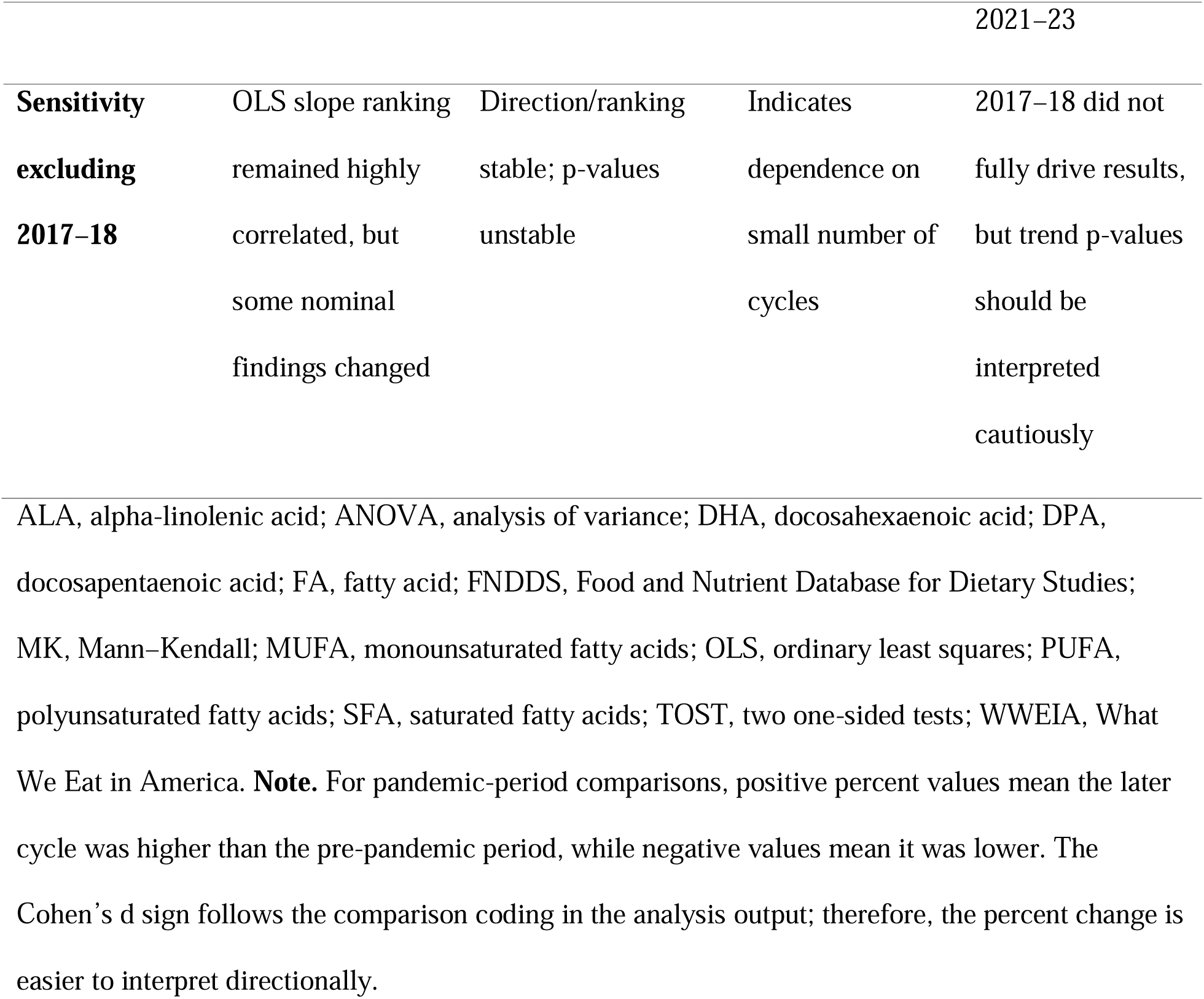
Summary of primary statistical findings and interpretation.

### 2.2 Data Processing

Raw FNDDS files were ingested in R (version 4.3) using schema-specific extraction routines, as three distinct structural formats were encountered across cycles: a wide-format nutrient table (2013-14, 2015-16), a long-format ingredient-nutrient-value file (2017-18, 2019-20), and a harmonized FoodData Central-aligned schema (2021-23) [14]. All outputs were standardized to a common long-format structure with fields for food code, WWEIA nutrient number, and per-100-g mean value. To limit leverage from extreme observations arising from concentrated specialty ingredients, flavor extracts, and highly fortified products, nutrient values were Winsorized at the 99th percentile within each analyte-cycle combination prior to computing cycle-level means [15]. Missing nutrient data were handled by restricting trend analyses to cycles with available values; analytes with valid data in fewer than four cycles were excluded from Mann-Kendall assessment. Because the number and definition of food codes varied across releases, especially in 2017-18, all cycle comparisons were interpreted as food-code-level database surveillance rather than as direct estimates of reformulation or intake.

### 2.3 Statistical Analysis

Temporal trends were assessed using three complementary approaches applied to cycle-level mean nutrient values for all 63 analytes. Ordinary least squares (OLS) regression modeled each Winsorized cycle mean as a linear function of cycle midpoint year, yielding annual slope estimates (β, in units per year), coefficient of determination (R²), and two-sided p-values. Because only five time points were available, OLS p-values were treated as exploratory. Multiple testing was addressed by Bonferroni correction for 63 simultaneous tests (α′ = 0.05/63 ≈ 0.00079); findings meeting this corrected threshold were designated as statistically confirmed trends, whereas unadjusted p < 0.05 findings were reported as nominal signals.

The Mann-Kendall test-a non-parametric rank correlation between nutrient values and time ranks, robust to non-normality and outliers-was applied to analytes with data in ≥4 cycles [16]. Welch’s one-way ANOVA was applied to food-code-level post-Winsorization data for each analyte across cycles to identify distributional heterogeneity [17]. ANOVA results were interpreted descriptively because food codes are not independent consumer observations and because differences may reflect database coverage, food-code definitions, fortification, or reformulation. Benjamini-Hochberg adjusted p-values were reported for distributional and pandemic-period comparisons.

Pandemic-period analyses compared pre-pandemic (FNDDS 2013-14 through 2017-18), pandemic-period (2019-20), and post-pandemic (2021-23) food-code composition. Pairwise Welch’s two-sample tests were derived from food-code-level summary statistics [18], with standardized differences reported as Cohen’s d [19]. Formal equivalence testing used the two-one-sided test (TOST) procedure with an equivalence bound of ±10% of the first comparison mean [20]. To avoid overinterpreting pre-pandemic/post-pandemic differences that could reflect the 2017-18 structural cycle, TOST was used only for the 2019-20 versus 2021-23 stability comparison. Sensitivity of OLS slopes to the 2017-18 cycle was assessed by repeating regressions on a four-point dataset excluding 2017-18 and comparing slopes with the five-cycle model. All analyses were conducted in R 4.3 using tidyverse, Kendall, and broom [14,21,22].

### 2.4 Ethical Considerations

All data used in this analysis are publicly available through USDA FoodData Central and required no primary data collection. No human subjects were enrolled, and no institutional review board approval was required. The NHANES dietary recall data underlying FNDDS construction were collected under protocols reviewed and approved by the National Center for Health Statistics Research Ethics Review Board, with written informed consent obtained from all NHANES participants.

## 3. Results

### 3.1 Nutrient Composition of the 2021-23 FNDDS Food-Code Universe

The 2021-23 FNDDS cycle included 5,431 food codes and provides the most recent food-code-level snapshot in this analysis. Per 100 g edible portion, the unweighted mean energy content was 197.8 kcal, with 20.77 g carbohydrate, 9.05 g total fat, 8.06 g protein, and 1.70 g dietary fiber. Sodium was 327.6 mg/100 g, while potassium, calcium, and magnesium were 204.0 mg, 70.5 mg, and 27.0 mg/100 g, respectively. Vitamin D content was 0.377 µg/100 g, while vitamin C was 5.26 mg/100 g. These values describe the unweighted FNDDS food-code universe and should not be interpreted as intake-weighted population exposure. Selected nutrient trajectories are shown in Figure 1, and complete cycle-level descriptive statistics are provided in Supplementary Table S1.

The 2021-23 fat profile showed 2.74 g/100 g saturated fatty acids, 3.19 g/100 g MUFA, and 2.15 g/100 g PUFA. Compared with 2013-14, total fat increased from 7.45 to 9.05 g/100 g, while PUFA increased from 1.68 to 2.15 g/100 g. However, these changes occurred alongside intermediate-cycle fluctuation, particularly the visibly elevated 2017-18 values for several macronutrients and fat measures. The main interpretation is therefore modest net compositional change with cycle-specific variation, not a simple monotonic increase across all energy-providing nutrients (Figure 1; Supplementary Table S1).

### 3.2 Pandemic-period composition

Pairwise pandemic-period analyses showed statistically detectable but small standardized differences when the pre-pandemic baseline was compared with either 2019-20 or 2021-23. Thirty-nine of 63 analytes differed after adjustment in the pre-pandemic versus 2019-20 comparison, and 39 of 63 differed after adjustment in the pre-pandemic versus 2021-23 comparison. However, the largest standardized difference remained below the conventional small-effect threshold (maximum |Cohen’s d| = 0.185 for pre-pandemic versus 2019-20 and 0.186 for pre-pandemic versus 2021-23; median |d| = 0.078 and 0.082, respectively). These results indicate statistically detectable food-code-level shifts with limited standardized magnitude (Figure 2; Supplementary Table S5).

**Figure 2.**
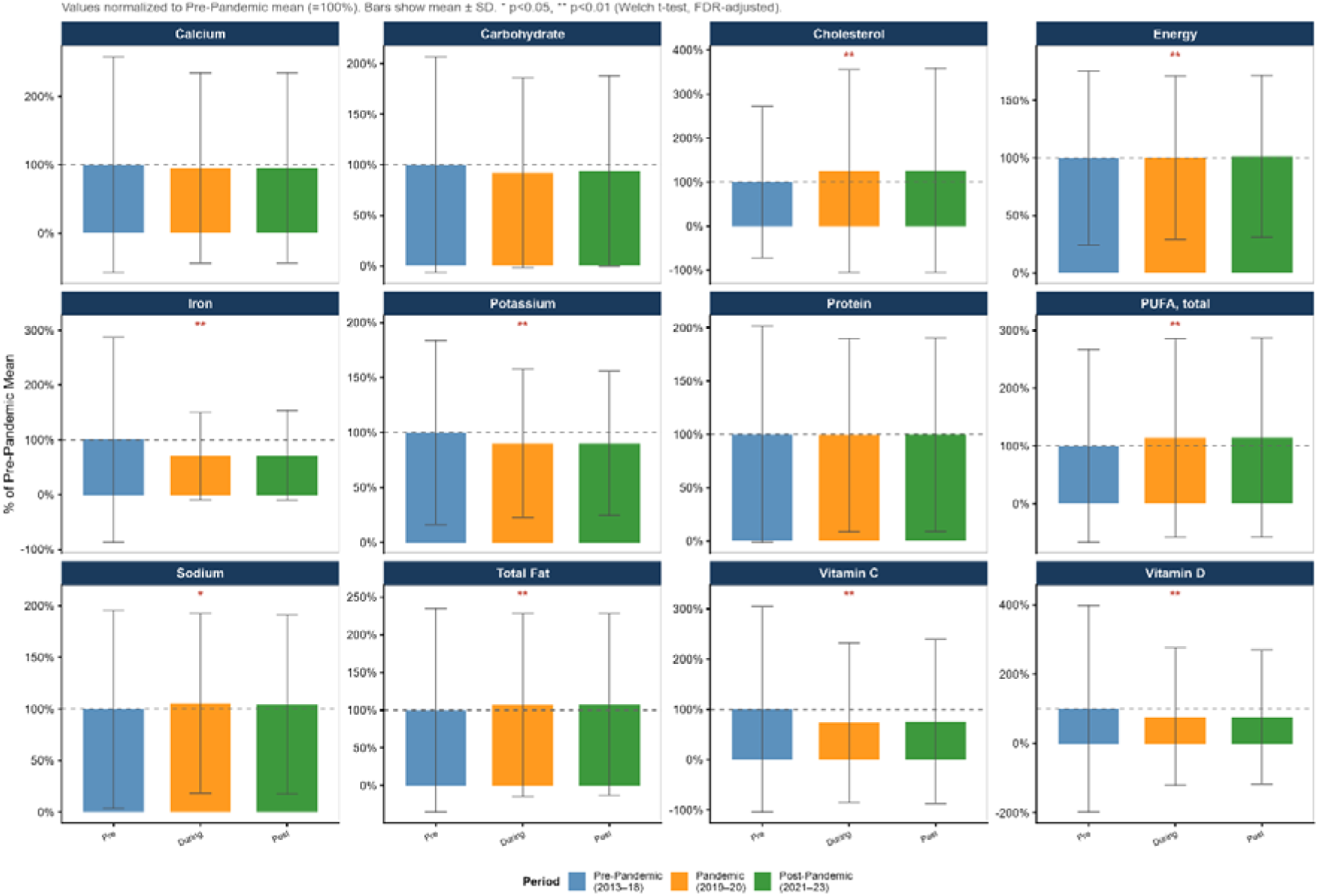
Pandemic-era shifts in nutrient composition: pre-pandemic, pandemic-period, and post-pandemic cycles.

In contrast, the direct comparison of 2019-20 with 2021-23 showed no statistically significant nutrient differences after adjustment, and standardized effects were negligible (maximum |Cohen’s d| = 0.026; median |d| = 0.007). TOST equivalence analysis found that 39 of 48 primary analytes were equivalent within ±10% bounds for the 2019-20 versus 2021-23 comparison. Thus, the most defensible pandemic-period interpretation is not that the full pre-pandemic and post-pandemic food-code universe was equivalent, but that the 2021-23 cycle was very similar to the immediately preceding 2019-20 cycle (Table 1; Figure 2; Supplementary Tables S5 and S9).

ALA, alpha-linolenic acid; ANOVA, analysis of variance; DHA, docosahexaenoic acid; DPA, docosapentaenoic acid; FA, fatty acid; FNDDS, Food and Nutrient Database for Dietary Studies; MK, Mann–Kendall; MUFA, monounsaturated fatty acids; OLS, ordinary least squares; PUFA, polyunsaturated fatty acids; SFA, saturated fatty acids; TOST, two one-sided tests; WWEIA, What We Eat in America. **Note.** For pandemic-period comparisons, positive percent values mean the later cycle was higher than the pre-pandemic period, while negative values mean it was lower. The Cohen’s d sign follows the comparison coding in the analysis output; therefore, the percent change is easier to interpret directionally.

### 3.3 Decade-scale effect sizes were small despite visible nutrient-specific changes

Across the full 2013-14 to 2021-23 interval, most nutrients had negligible or small Cohen’s d values. The largest positive effect sizes were observed for energy, linoleic acid, total fat, total PUFA, palmitic acid, oleic acid, and total MUFA. The largest negative effect sizes were observed for lutein + zeaxanthin, added vitamin B12, vitamin A, DHA, beta-carotene, water, arachidonic acid, vitamin K, total vitamin B12, and vitamin B6. The largest absolute effect size was 0.179, supporting the interpretation that decade-scale food-code-level compositional differences were modest in standardized terms (Figure 3; Supplementary Table S6).

**Figure 3.**
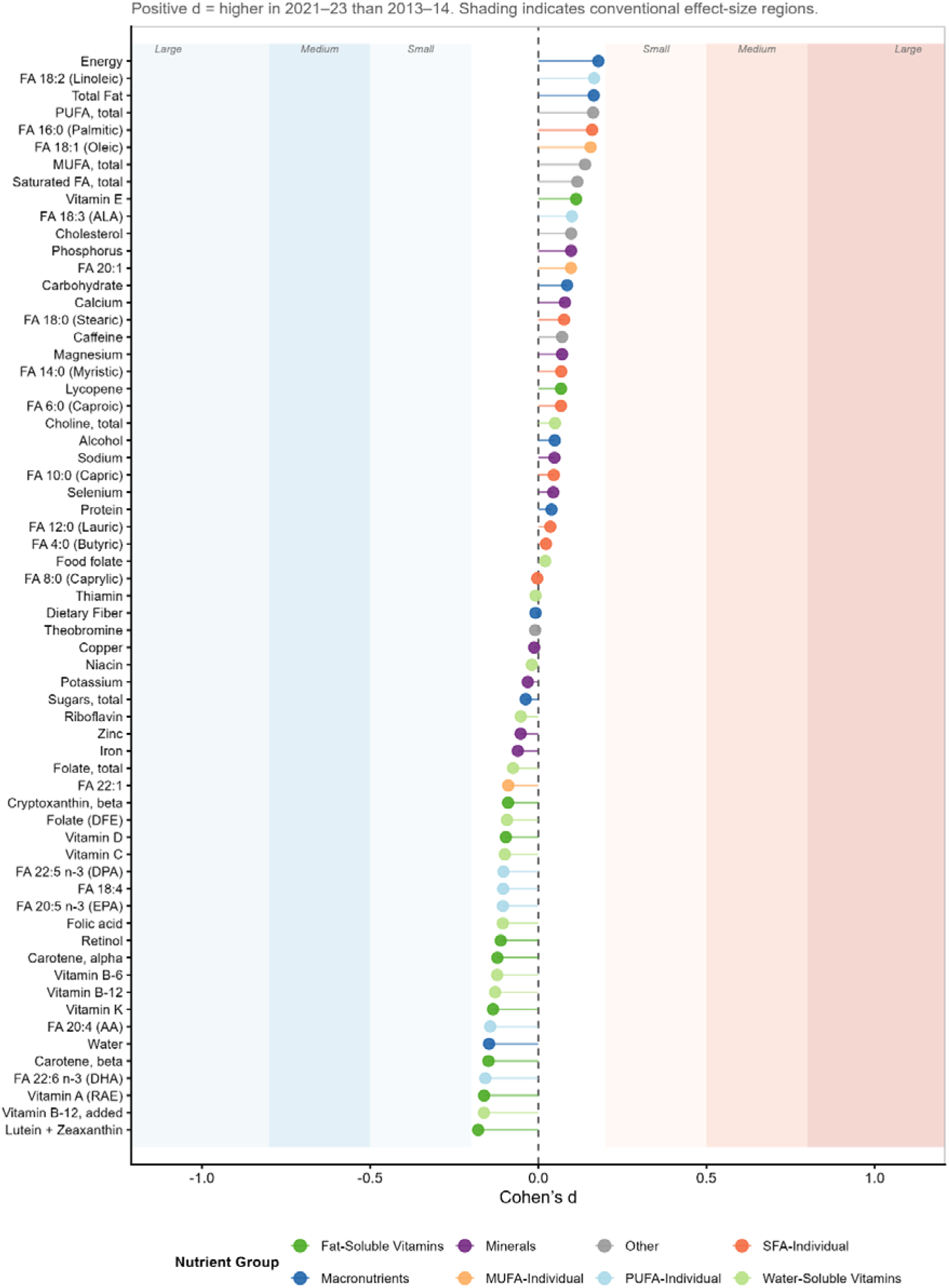
Effect sizes for nutrient composition change between 2013–14 and 2021–23.

The effect-size pattern also shows why percent change alone can be misleading. Some nutrients with large relative percentage changes, especially low-concentration fatty acids and carotenoids, had small absolute changes and small standardized effects. Conversely, macronutrients and common fatty acids showed smaller proportional shifts but contributed more clearly to the overall compositional pattern because they occur at higher absolute concentrations (Figure 3; Supplementary Table S6).

### 3.4 Fat-quality changes were the clearest exploratory nutrient-specific signal

The most coherent exploratory nutrient-specific pattern was a shift in fatty acid composition. Total PUFA increased from 1.68 g/100 g in 2013-14 to 2.15 g/100 g in 2021-23. Linoleic acid increased from 1.46 to 1.90 g/100 g, and alpha-linolenic acid increased from 0.151 to 0.176 g/100 g. In contrast, long-chain omega-3 fatty acids decreased over the same period: EPA decreased from 0.0079 to 0.0050 g/100 g, DPA from 0.0037 to 0.0026 g/100 g, and DHA from 0.0156 to 0.0074 g/100 g. Thus, any increase in total PUFA appears to be driven mainly by plant-oil-associated PUFA rather than marine omega-3 fatty acids (Figure 4A; Supplementary Figure S4; Supplementary Table S1).

**Figure 4.**
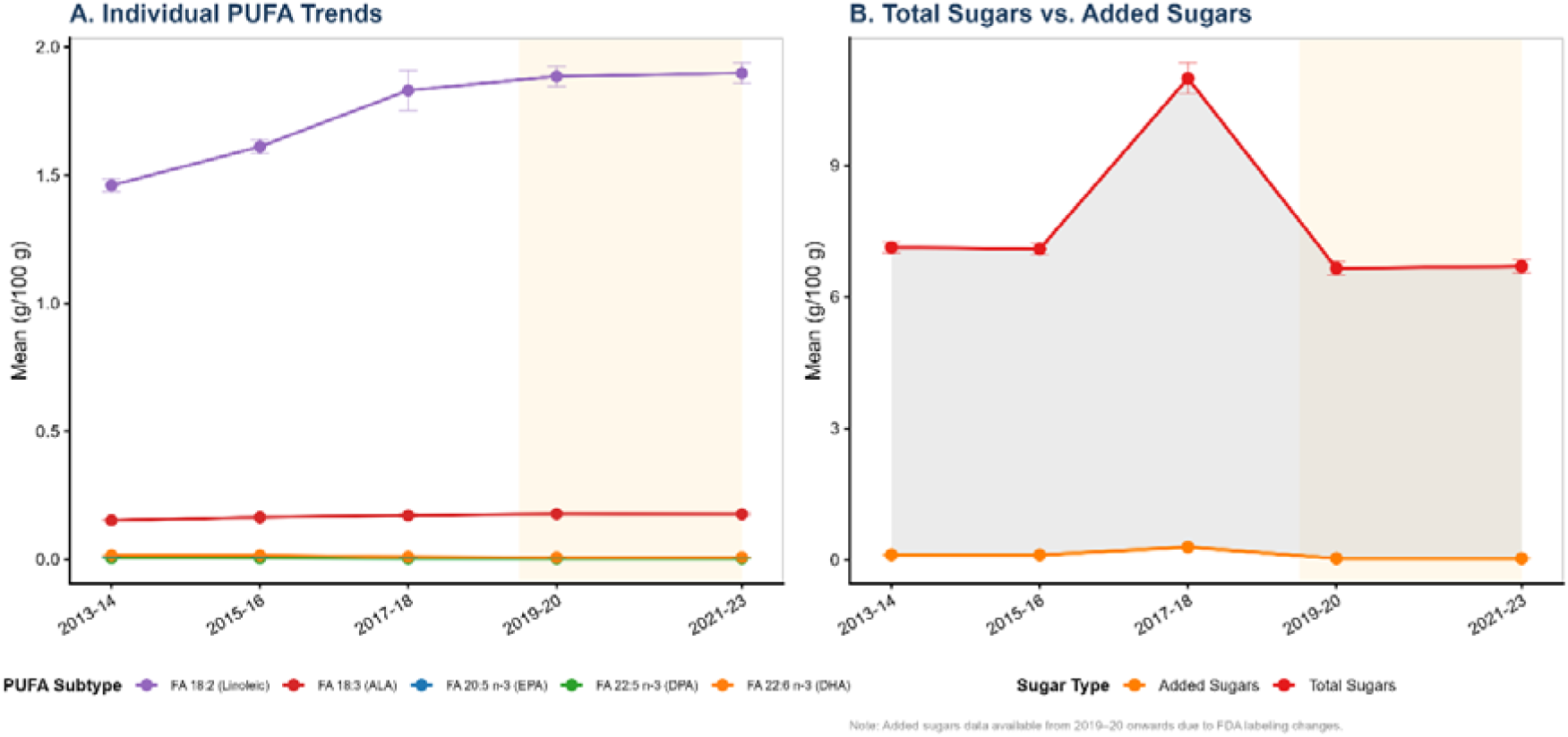
Polyunsaturated fatty acid subtypes and sugar profile trends, 2013–2023.

Total sugars showed a marked peak in 2017-18, followed by lower values in 2019-20 and 2021-23. Total sugars were 7.13 g/100 g in 2013-14, 10.99 g/100 g in 2017-18, 6.65 g/100 g in 2019-20, and 6.70 g/100 g in 2021-23. Because comparable added-sugar data were not available for the full study window, added sugars should not be interpreted as a decade-long trend. Instead, the sugar analysis supports a narrower conclusion: total sugar density did not show a sustained increase by 2021-23, and added-sugar interpretation remains constrained by data availability (Figure 4B; Supplementary Table S1).

### 3.5 Linear trend testing identified nominal signals but no Bonferroni-confirmed trend

OLS regression across the five cycle midpoints identified eight nominally significant linear trends at p < 0.05. These were beta-cryptoxanthin, linoleic acid, FA 20:1, DHA, alpha-linolenic acid, total PUFA, DPA, and FA 18:4. Total PUFA showed a positive nominal trend, increasing by 0.057 g/100 g per year (p = 0.028), while linoleic acid increased by 0.054 g/100 g per year (p = 0.024). However, none of the 63 OLS trend tests remained statistically significant after Bonferroni correction. These findings should therefore be presented as exploratory or nominal trend signals rather than confirmed corrected associations (Table 1; Supplementary Figure S5; Supplementary Table S2).

The Mann–Kendall analysis supported the same general interpretation. Total PUFA and linoleic acid were nominally significant by Mann–Kendall testing, but neither survived multiple-testing adjustment. This reinforces the conclusion that the main decade-scale signal is directionally consistent and biologically interpretable, but statistically modest given the short five-cycle time series (**Supplementary Table S3**).

### 3.6 Distributional differences were widespread across cycles

Although few nutrients showed robust monotonic linear trends, Welch’s ANOVA detected broad distributional differences across FNDDS cycles. At the nominal p < 0.05 level, 61 of 63 nutrients differed across cycles; after adjustment, 57 of 63 remained significant. The strongest distributional differences were observed for water, added vitamin B12, energy, lutein + zeaxanthin, total vitamin B12, beta-carotene, vitamin B6, vitamin A, DHA, carbohydrate, total fat, iron, folic acid, and palmitic acid. These results indicate that the food-code-level nutrient distributions changed across cycles even when average linear trends were not statistically confirmed (Table 1; Supplementary Figure S2; Supplementary Table S4).

This distinction is important for interpretation. The FNDDS food-code universe did not show many simple corrected linear mean trends, but its nutrient distributions were not static. Reformulation, changes in food-code coverage, fortification patterns, analytical updates, and shifts in the types of foods represented in FNDDS may all alter distributions without producing a strong monotonic mean trend. Therefore, ANOVA results should be interpreted as evidence of broad database-level compositional heterogeneity, not as evidence that all nutrients changed in one consistent direction (Supplementary Figure S2; Supplementary Table S4).

### 3.7 Sensitivity analysis showed strong slope stability despite nominal-status changes

The 2017-18 cycle showed a noticeable increase in several analytes, including energy, carbohydrate, total sugars, total fat, and dietary fiber, and also had a substantially smaller number of food codes than adjacent cycles. To assess whether this cycle had undue influence, OLS models were repeated after excluding 2017-18. Slope estimates from the four-cycle and five-cycle models were highly correlated, indicating that broad trend direction and relative ranking were stable. However, nominal significance status changed for several nutrients because the reduced model altered standard errors and only four time points remained (Supplementary Figure S3; Supplementary Table S7).

This sensitivity analysis supports two conclusions. First, the broad direction of estimated nutrient change was not driven solely by 2017-18. Second, nominal p-values should be interpreted with caution because the time series is short and sensitive to individual cycles. The most defensible interpretation is therefore based on consistency across figures, effect sizes, and sensitivity results rather than on any single p-value threshold (Table 1; Supplementary Figure S3; Supplementary Table S7).

## 4. Discussion

The 2021-23 FNDDS cycle provides the most current food-code-level characterization in this analysis. The unweighted sodium content of 327.6 mg/100 g, while essentially unchanged from the preceding cycle (331.4 mg/100 g in 2019-20), remains notable in the context of the Dietary Guidelines sodium limit of 2,300 mg/day [3,23]. Dietary fiber (1.70 g/100 g) and key shortfall micronutrients, including vitamin D (0.377 µg/100 g) and potassium (204.0 mg/100 g), also remained low in the unweighted food-code summaries relative to public health concerns about U.S. dietary adequacy [3,24]. Because this study did not apply NHANES consumption weights, these values should be interpreted as database-level composition, not direct population intake. Nevertheless, they identify nutrient domains where future intake-weighted surveillance remains important.

The widespread distributional heterogeneity detected by Welch’s ANOVA (61 of 63 analytes at nominal p < 0.05; 57 after adjustment) is an important methodological finding, but it should not be overinterpreted as direct proof of industry-wide reformulation. Distributional shifts in FNDDS can arise from multiple mechanisms: true reformulation, fortification changes, changes in food-code coverage, altered recipe assumptions, schema harmonization, or differences in the types of foods represented across cycles. This explains the dissociation between widespread ANOVA findings and limited corrected OLS trends. Future FNDDS surveillance should therefore combine mean-level trend analysis with distributional summaries and should document database structural changes explicitly [25,26].

The clearest biologically interpretable exploratory signal was the increase in total PUFA and linoleic acid, but this signal did not survive correction for 63 simultaneous OLS tests and should be treated as hypothesis-generating. Its direction is nevertheless consistent with the fat-quality emphasis in U.S. dietary guidance [3,27], the industrial removal of partially hydrogenated oils following the FDA’s GRAS determination and compliance period [1], and broader reformulation incentives related to saturated fat and oil substitution [28]. The subtype pattern suggests that the increase was driven mainly by plant-oil-associated PUFA rather than marine omega-3 enrichment; EPA and DHA decreased in the food-code means across the study window, despite consumer and clinical interest in marine omega-3 fatty acids [29].

The pandemic-period findings require careful wording. Conventional significance testing showed adjusted differences between the pre-pandemic baseline and both 2019-20 and 2021-23, but all standardized effects were small. Conversely, the direct 2019-20 versus 2021-23 comparison showed no adjusted differences, negligible effect sizes, and formal equivalence within ±10% bounds for 39 of 48 primary analytes (Table 1; Supplementary Tables S5 and S9) [30]. Therefore, this study supports post-2019-20 food-code-level stability rather than strong evidence that the entire pre-pandemic and post-pandemic food universe was unchanged. This interpretation is consistent with the literature describing rapid adaptation and partial normalization of food supply chains following the first pandemic shock [4,31].

Our findings also help reconcile aggregate database stability with targeted studies showing nutrient changes in specific food categories. Category-level audits of packaged foods have reported sodium reductions following voluntary reduction initiatives and sugar reformulations in some product categories [8,9,32]. Such localized changes may be attenuated when averaged across thousands of FNDDS food codes, especially when some categories move in opposite directions or when database coverage changes across cycles [33]. Because this analysis was not intake-weighted, the findings should not be interpreted as changes in actual U.S. dietary intake. Population dietary exposure requires individual-level NHANES intake data, and prior studies have shown that U.S. adult dietary intake and diet quality changed over time in ways that cannot be inferred from food-code composition alone [34,35]. The present study, therefore, should be viewed as a broad surveillance screen that can generate hypotheses for category-specific analyses, not as a substitute for product-level or intake-weighted studies.

Several limitations require consideration. First, analyses were unweighted by consumption volume, so rare food codes and commonly consumed staples contributed equally; future analyses should integrate FNDDS nutrient values with NHANES dietary weights to estimate intake-weighted exposure. Second, FNDDS cycles differed in schema, food-code definitions, and number of records, particularly in 2017-18, so observed changes may partly reflect database structure rather than true food-supply reformulation. Third, food-code-level Welch tests treat records as analytic observations and may produce very small p-values when sample sizes are large; effect sizes and sensitivity analyses are therefore more informative than statistical significance alone. Fourth, added sugars lacked full-window comparability, limiting conclusions about sugar reformulation. Fifth, the five-cycle OLS time series has limited power and should be interpreted as exploratory despite multiplicity correction.

## 5. Conclusions

The unweighted FNDDS food-code universe showed broad distributional heterogeneity across 2013-2023, but no nutrient had a Bonferroni-confirmed decade-long linear trend. Total PUFA and linoleic acid were the most coherent nominal signals and may warrant targeted follow-up, but they should not be presented as corrected findings. Pandemic-period analyses showed small standardized differences relative to the pre-pandemic baseline, while the 2021-23 cycle was highly similar to 2019-20, including formal ±10% equivalence for 39 of 48 primary analytes (Table 1; Figure 2; Supplementary Table S9). The 2021-23 FNDDS cycle provides a useful current database-level baseline for future nutrition surveillance, but intake-weighted analyses are needed before inferring population dietary exposure or progress toward public health targets (Figure 1). Continued multi-cycle surveillance, paired with transparent documentation of database structural changes, is essential for detecting subtle but policy-relevant nutrient composition patterns.

## Declarations

### Ethics approval and consent to participate

Not applicable. This study did not involve human participants, patient data, animals, or biological specimens requiring ethical approval.

### Consent for publication

Not applicable.

### Availability of data and materials

The primary data used in this study is avaalible through https://fdc.nal.usda.gov/download-datasets. All data generated or analyzed during this study are included in this published article and its supplementary materials. Further information may be available from the corresponding author upon reasonable request.

### Competing interests

The authors declare that they have no competing interests.

### Funding

The authors received no specific financial support, grant, or funding for this research, authorship, and/or publication of this article.

### Authors’ contributions

**O.I.M.** contributed to methodology, data curation, formal analysis, and drafted the original manuscript.

**M.E.A.** conceptualized the study, contributed to study design, methodology, supervision, validation, project administration, and critical revision of the manuscript. **B.M.H.** contributed to the literature review, data collection, investigation, analysis, and manuscript review and editing. **A.Z.A.** contributed to data interpretation, visualization, manuscript review, editing, and revision. **A.S.** contributed to investigation, validation, analytical support, and manuscript editing. All authors read, reviewed, and approved the final manuscript.

All authors read and approved the final manuscript.

## Acknowledgments

Not applicable.

## Notes

### Competing Interest Statement

The authors have declared no competing interest.

